# Psychological antecedents towards COVID-19 vaccination using the Arabic 5C validated tool: An online study in 13 Arab countries

**DOI:** 10.1101/2021.08.31.21262917

**Authors:** Marwa Shawky Abdou, Khalid A. Kheirallah, Maged Ossama Aly, Ahmed Ramadan, Yasir Ahmed Mohammed Elhadi, Iffat Elbarazi, Ehsan Akram Deghidy, Haider M. El Saeh, Karem Mohamed Salem, Ramy Mohamed Ghazy

## Abstract

**Background and aim:** Following emergency approval of vaccines, the amount of scientific literature investigating population hesitancy towards vaccination against the novel coronavirus disease (COVID-19) has increased exponentially. Nevertheless, the associated psychological behaviors with this phenomenon are still not clearly understood. This study aims to assess the psychological antecedents of the Arab population toward COVID-19 vaccines.

**Methods:** A cross-sectional, online study using a validated Arabic version of the 5C questionnaire was conducted through different media platforms in different Arabic-speaking countries. The questionnaire included three sections: socio-demographics, COVID-19 related questions, and the 5C scale of vaccine psychological antecedents, namely confidence, complacency, constraints, calculation, and collective responsibility.

**Results:** A total of 4,474 participants, 40.8% males from 13 Arab countries were included in the study. About 26.7% of participants had confidence in COVID-19 vaccination, 10.7% had complacency, 96.5% had no constraints, 48.8% had calculation and 40.4% had collective responsibility. The 5C antecedents showed variation among countries with confidence and collective responsibility being higher in the United Arab Emirates (UAE) (59% and 58%, respectively), complacency and constraints were higher in Morocco (21% and 7%, respectively) and calculation was higher in Sudan (60%). Regression analysis revealed that sex, age, educational degrees, being a health care professional, getting a COVID-19 infection, having a relative infected or died from COVID-19 can affect the 5C psychological antecedents by different degrees.

**Conclusion and recommendations:** Wide variations of psychological antecedents between Arab countries exist. Different determinants can affect vaccine psychological antecedents.

## Introduction

Coronavirus disease (COVID-19) was initially identified in China in December 2019. Since then, infection with the Severe Acute Respiratory Syndrome Coronavirus 2 (SARS-CoV-2) has swamped the globe. ^(1)^ The global threat of the pandemic is still on the rise with more than 200 million cases and more than 4 million deaths. ^(2)^ The situational report of the World Health Organization (WHO) showed that the Eastern Mediterranean Region (EMR) ranks fourth in the number of COVID-19 cases, totaling more than 12 million cases and nearly 240 thousand deaths. ^(3-5)^ Among the Arab world, as of 27 July 2021, Iraq had disclosed the highest number of cases followed by Jordan, the United Arab Emirates (UAE), Morocco, and Tunisia. Associated deaths were highest in Tunisia, Iraq, and Egypt. ^(5)^ The new variant strains, like the delta strain, are posing another threat to these countries and call for immediate action in terms of precautionary measures including vaccination. ^(6)^

As the Non-Pharmaceutical Intervention (NPI) measures against COVID-19, such as social distancing and curfew, were not enough to mitigate the spread of the virus, vaccination became the vital measure against the threats associated with COVID-19. ^(7)^ There is a global consensus supporting the notion that COVID-19 vaccines are likely the most effective approach to control the pandemic. ^(8)^ The unprecedented research efforts, and global coordination, have resulted in the rapid development and administration of vaccines likely to control COVID-19. ^(9)^ Since the emergence of COVID-19, there has been a surge in vaccines development. By 24 September 2020, a staggering number of vaccines had been under pre-clinical development, of which 43 had entered clinical trials, including some approaches that have not previously been licensed for human vaccines. ^(10)^

Strenuous scientific efforts against the pandemic have led to the utilization of different modalities and novel techniques as vaccine platforms. COVID-19 vaccines are either mRNA (manufactured by Moderna and BioNTech/Pfizer), inactivated virus (Sinovac, Sinopharm), viral vector (Oxford/AstraZeneca, Gamaleya, Janssen/Johnson & Johnson, CanSino), or protein sub-unit (Novavax). The vaccine produced by BioNTech/Pfizer has been deployed to the public as the first-ever licensed COVID-19 vaccine. ^(11)^ More countries are entering the race of vaccine development such as Cuba, Brazil, and others. ^(12)^

People all over the globe have developed concerns regarding the COVID-19 authorized vaccines due to many reasons. ^(10)^ Some of these reasons include the quick development and release of the vaccines, the conspiracy theory including the origin of the vaccine, and possibly due to other factors to be explored. Vaccine Hesitancy (VH) existed before the COVID-19 emergence which augmented people’s doubts and pushed people to become against vaccination. ^(13)^ One of the major obstacles against the success of the vaccination programs is VH, which can affect both the individual, by having a greater risk to get infected, and the community, by easily transmitting the pathogen. ^(14)^ As such, VH is among nine other health challenges considered as global health threats by the World Health Organization (WHO) in 2019. ^(15)^ VH is defined as a behavior associated with delay in acceptance or refusal of vaccines despite available services. It is a complex and context-specific behavior that varies across time, place, and disease but is still influenced by factors such as complacency, convenience, and confidence. ^(16)^

The Five psychological antecedents of the vaccination tool (5C model) were developed by Betsch and colleagues. ^(17)^ The 5C model identifies psychological antecedents for everyone that may indicate whether an individual will vaccinate or not. These five antecedents are confidence, complacency, constraints, calculation, and collective responsibility. The 5C scale is used to assess these 5 psychological antecedents of vaccination and to provide insights into how the individual may think, feel, and behave regarding vaccination. These antecedents impact the vaccination behavior to varying degrees and assess the mental portrayals, attitudinal and behavioral propensities that result from the environment and context the individual lives in. ^(17-19)^ These antecedents are used nowadays as a framework to assess VH in the high-income countries to find whether people will or will not take the COVID-19 vaccine. ^(20)^

The COVID-19 VH rate was reported to be significantly different by socio-demographic characteristics, seasonal flu vaccination status, COVID-19 risk perception, and perceived benefits and clinical barriers of the COVID-19 vaccine. ^(21)^ In Hong Kong, 63% of nursing staff were likely to take the COVID-19 vaccine when available. ^(22)^ In low- and middle-income countries, the vaccine acceptance rate ranged from 66.5% in Burkina Faso to 96.6% in Nepal with an overall acceptance rate of 80.3%. ^(23)^ At this stage of the pandemic, especially as vaccine compliance remains variable and inconsistent, public health officers and policymakers, especially in developing countries where healthcare resources are limited, need to understand the reasons and factors associated with VH. To investigate the associated physiological behaviors with this phenomenon, this study has been conceived, the present study aimed to investigate the psychological antecedents of the Arab population towards COVID-19 vaccination.

## Method

### Study design, sampling, and data collection

A cross-sectional, web-based, anonymous survey using the Arabic-validated version of the 5C questionnaire ^(24)^ was conducted between December 2020 and February 2021. The research team used social media platforms such as Facebook, Twitter, and WhatsApp, to recruit potential participants.

### Data collection tool

The survey consists of the following sections: The first section includes the sociodemographic characteristics (age, sex, residence, level of education, marital status, occupation, and presence of comorbidities). The second section assesses past COVID-19 infection and vaccination history (previous infection, family history, mortality, influenza vaccination, types of COVID-19 vaccines, web information of COVID-19 vaccine). In the third section, there are 15 questions covering the five domains of the 5C investigated. These are confidence, complacency, constraints, calculation, and collective responsibility. Each domain contains 3 questions with a 7-point Likert scale (1 to 7). The cutoff point of confidence, complacency, constraints, calculation, and collective responsibility were 5.7, 4.7, 6.0, 6.3, and 6.2, respectively. ^(25)^

### Statistical analysis

The collected data was wrangled, coded, and analyzed using Python 3.9.2 software. The quantitative variables were expressed using mean ± SD, whereas counts (percentages) were utilized to describe the categorical variables. The Chi-square test was used to estimate pairwise correlations between categorical variables. Finally, respondents were categorized (Yes/No) based on their mean 5C scores with reference to the cutoff points determined before. Further, five stepwise binary logistic regression models were performed to estimate significant predictors for confidence, complacency, calculation, constraints, and collective responsibility. Coefficients and correlations with *P-*value <0.05 were considered statistically significant.

### Ethical considerations

The study was approved by the Ethics Committee of the Faculty of Medicine, Alexandria University, Egypt (IRB No: 00012098). The researchers complied with the International Guidelines for Research Ethics. ^(26)^ Participants were informed that their participation was voluntary, and consent was obtained before administering the survey.

## Results

### Characteristics of respondents

A total of 4,474 participants from 13 Arabic countries were included in the current analysis, of which, 40.8% were males. The mean (SD) age was 32.48 ±10.76 years. The majority were either married (50.7%) or single (44.4%). About half of participants had a university degree (50.3%), the majority reported no chronic illnesses (82.7%), about two-fifth (40%) were health care professionals (HCPs), more than one-quarter (27.9%) reported being vaccinated against COVID-19 infection, nearly half (47.5%) had at least one person who got infected with COVID-19, about one third (33.6%) reported at least one relative died from COVID-19, and only 21.5% knew that there are different types of COVID-19 vaccines (Table 1).

**Table 1;.** Characteristics of respondents

| **Variables** | **No.** | **%** |
| --- | --- | --- |
| **Sex** |  |  |
| **Male** | 1,825 | 40.8 |
| **Female** | 2,649 | 59.2 |
| **Age (years) Mean ± SD** | 32.48 ± 10.76 | |
| **Marital status** |  |  |
| **Single** | 1,804 | 44.4 |
| **Married** | 2,058 | 50.7 |
| **Divorced** | 139 | 3.4 |
| **Widow** | 60 | 1.5 |
| **Educational status** |  |  |
| **Pre-university** | 323 | 7.2 |
| **Technical/ vocational education** | 129 | 2.9 |
| **University degree** | 2,243 | 50.3 |
| **Postgraduate degree** | 1,502 | 33.7 |
| **Other** | 266 | 6.0 |
| **Having a Chronic disease** | 773 | 17.3 |
| **Being HCP** | 1,789 | 40.0 |
| **Previously infected with COVID-19** | 1,025 | 27.9 |
| **Relative infected with COVID-19** | 1,915 | 47.5 |
| **Relatives died from COVID-19** | 1,390 | 33.6 |
| **Getting the flu vaccine** | 104 | 26.1 |
| **Knowing different COVID-19 vaccines** | 3510 | 78.5 |
| **Best COVID-19 vaccine** |  |  |
| • **Moderna** | 24 | 7.3 |
| • **Pfizer- BioNTech** | 194 | 58.8 |
| • **Oxford-AstraZeneca** | 52 | 15.8 |
| • **Sinopharm** | 51 | 15.5 |
| • **Sputnik V** | 9 | 2.7 |
| **Infected with COVID-19 after vaccination** | 48 | 21.4 |
| **Knowing COVID-19 vaccine Instructions** | 125 | 55.8 |
| **Internet search about COVID-19 vaccine** | 135 | 60.3 |
| **Getting COVID vaccine if free** | 140 | 62.5 |

### Psychological antecedents of vaccination among respondents

Figure 1 shows that 26.7% of participants had confidence in COVID-19 vaccination, 10.7% had complacency, 96.5% had no constraints, 48.8% had calculation and 40.4% had collective responsibility.

**Figure 1:**
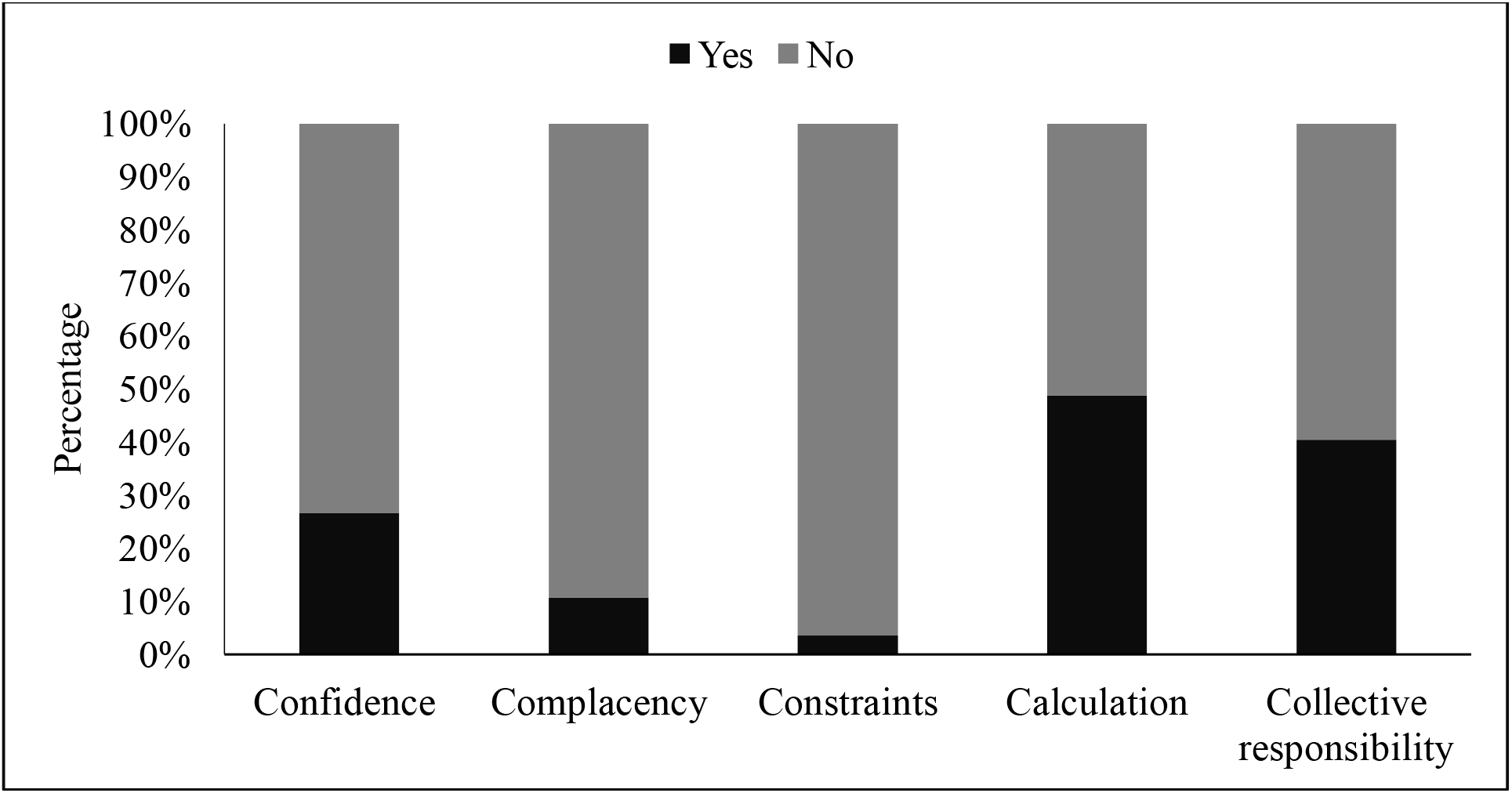
Psychological antecedents of vaccination among respondents

### Psychological antecedents to COVID-19 vaccination among the studied countries

As shown in (Figure 2), confidence and collective responsibility were higher among the UAE population (59% and 58%, respectively), complacency and constraints were higher among the Morocco population (21% and 7%, respectively), while calculation was higher among Sudanese (60%). On the other hand, Egypt had the lowest confidence (15%), Lebanon had the lowest complacency (7.5%), Sudan had the lowest constraint (1.2%), Iraq had the lowest calculation (36%) and Morocco had the lowest collective responsibility (25%)

**Figure 2:**
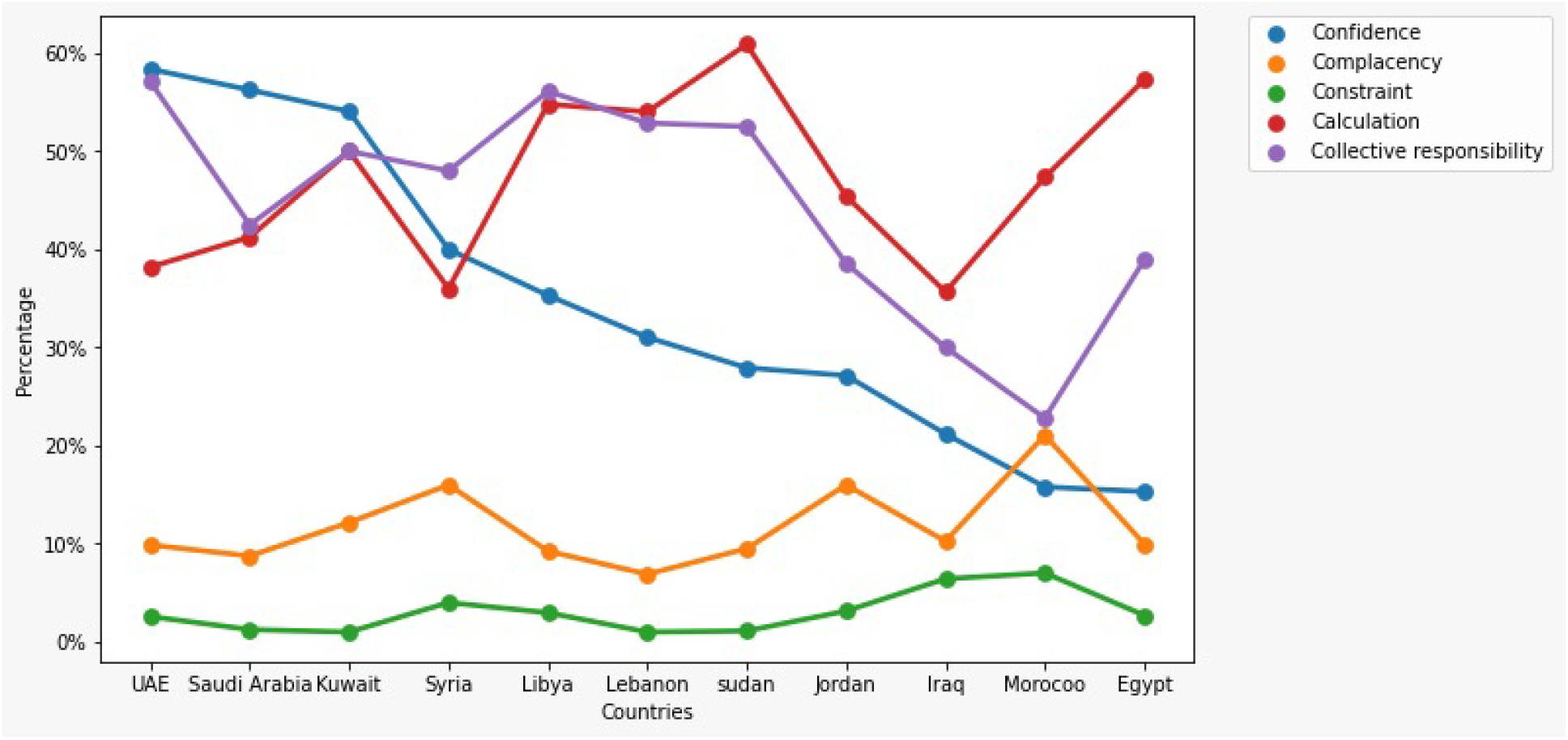
Psychological antecedents to COVID-19 vaccination among the studied countries

### Bi-variate analysis of the 5C domains and the independent variables

Table 2 shows the distribution of each of the 5C domains by independent variables at the bi-variate levels.

**Table 2:** Distribution of each of the 5C domains by independent variables at the bi-variate levels.

| Variable | Confidence<br>n(%) |  | Complacency<br>n(%) |  | Constraints<br>n(%) |  | Calculation<br>n(%) |  | Responsibility<br>n(%) |  |
| --- | --- | --- | --- | --- | --- | --- | --- | --- | --- | --- |
|  | Yes | No | Yes | No | Yes | No | Yes | No | Yes | No |
| **Sex** |  |  |  |  |  |  |  |  |  |  |
| **Male** | 560<br>(30.68) | 1265<br>(69.32) | 209<br>(11.45) | 1616<br>(88.55) | 58<br>(3.18) | 1767<br>(96.82) | 817<br>(44.77) | 1008<br>(55.23) | 746<br>(40.88) | 1079<br>(59.12) |
| **Female** | 637<br>(24.05) | 2012<br>(75.95) | 268<br>(10.18) | 2381<br>(89.88) | 98<br>(3.70) | 2551<br>(96.30) | 1368<br>(51.64) | 1281<br>(48.36) | 1064<br>(40.17) | 1585<br>(59.83) |
| *P-value* | *<0.001* |  | *0.169* |  | *0.394* |  | *0.007* |  | *0.656* |  |
| **Marital status** |  |  |  |  |  |  |  |  |  |  |
| **Single** | 453<br>(25.11) | 1351<br>(74.89) | 185<br>(10.25) | 1619<br>(89.75) | 70<br>(3.88) | 1734<br>(96.12) | 832<br>(45.62) | 972<br>(54.38) | 733<br>(40.63) | 1071<br>(59.37) |
| **Married** | 545<br>(26.48) | 1513<br>(73.52) | 225<br>(10.93) | 1833<br>(89.07) | 68<br>(3.30) | 1990<br>(96.70) | 1037<br>(50.39) | 1021<br>(49.61) | 838<br>(40.72) | 1220<br>(59.28) |
| **Divorced** | 35<br>(25.18) | 104<br>(74.82) | 15<br>(10.79) | 124<br>(89.21) | 5<br>(3.60) | 134<br>(96.40) | 57<br>(41.01) | 82<br>(58.99) | 43<br>(30.94) | 96<br>(69.06) |
| **Widow** | 27(45) | 33(55) | 4(6.67) | 56<br>(93.33) | 1(1.67) | 59<br>(98.33) | 33(55) | 27(45) | 25<br>(41.67) | 35<br>(58.33) |
| *P-value* | *0.006* |  | *0.693* |  | *0.668* |  | *0.011* |  | *0.148* |  |
| **Educational status** |  |  |  |  |  |  |  |  |  |  |
| **Pre-university** | 108<br>(33.44) | 215<br>(66.56) | 46<br>(14.24) | 277<br>(85.76) | 9(2.79) | 314<br>(97.21) | 138<br>(42.72) | 185<br>(57.28) | 129<br>(39.94) | 194<br>(60.06) |
| **Technical/ vocational education** | 45<br>(34.88) | 84<br>(65.12) | 16<br>(12.40) | 113<br>(87.60) | 5(3.88) | 124<br>(96.12) | 50<br>(38.76) | 79<br>(61.24) | 53<br>(41.09) | 76<br>(58.91) |
| **University degree** | 581<br>(25.90) | 1662<br>(74.10) | 269<br>(11.99) | 1974<br>(88.01) | 83<br>(3.70) | 2160<br>(96.30) | 1063<br>(47.39) | 1180<br>(52.61) | 904<br>(40.30) | 1339<br>(59.70) |
| **Postgraduate degree** | 391<br>(26.03) | 1111<br>(73.97) | 119<br>(7.92) | 1383<br>(92.08) | 43<br>(2.86) | 1459<br>(97.14) | 839<br>(55.86) | 663<br>(44.14) | 648<br>(43.14) | 854<br>(56.86) |
| **Other** | 71<br>(26.69) | 195<br>(73.31) | 27<br>(10.15) | 239<br>(89.85) | 16<br>(6.02) | 250<br>(93.98) | 93<br>(34.96) | 173<br>(65.04) | 75<br>(28.20) | 191<br>(71.80) |
| *P-value* | *0.003* |  | *0.011* |  | *0.107* |  | *<0.001* |  | *<0.001* |  |
| **Being a HCP** | 459<br>(25.66) | 1330<br>(74.34) | 135<br>(7.55) | 1654<br>(92.45) | 45<br>(2.52) | 1744<br>(97.48) | 956<br>(53.44) | 833<br>(46.56) | 815<br>(45.56) | 974<br>(54.44) |
| *P-value* | *0.187* |  | *<0.001* |  | *0.004* |  | *<0.001* |  | *<0.001* |  |
| **Get COVID** | 198<br>(19.32) | 827<br>(80.68) | 126<br>(12.29) | 899<br>(87.71) | 55<br>(5.37) | 970<br>(94.63) | 454<br>(44.29) | 571<br>(55.70) | 357<br>(34.83) | 668<br>(65.17) |
| *P-value* | *<0.001* |  | *0.0341* |  | *0.001* |  | *0.069* |  | *<0.001* |  |
| **Relative infected with COVID** | 503<br>(26.27) | 1412<br>(73.73) | 187<br>(9.77) | 1728<br>(90.23) | 70<br>(3.67) | 1845<br>(96.34) | 938<br>(48.98) | 977<br>(51.02) | 806<br>(42.09) | 1109<br>(57.91) |
| *P-value* | *0.06* |  | *0.095* |  | *0.392* |  | *0.361* |  | *0.102* |  |
| **Relative died from COVID** | 342<br>(24.60) | 1048<br>(75.40) | 146<br>(10.50) | 1244<br>(89.50) | 52<br>(3.74) | 1338<br>(96.26) | 741<br>(53.31) | 649<br>(46.69) | 584<br>(39.42) | 806<br>(60.58) |
| *P-value* | *0.033* |  | *0.937* |  | *0.316* |  | *0.002* |  | *0.395* |  |
| **Getting Flu vaccine** | 35<br>(33.65) | 69<br>(66.35) | 10<br>(9.62) | 94<br>(90.38) | 5<br>(4.81) | 99<br>(95.19) | 52 (50) | 52 (50) | 42<br>(40.38) | 62<br>(59.62) |
| *P-value* | *0.007* |  | *0.587* |  | *0.911* |  | *0.252* |  | *0.049* |  |
| **Knowing different COVID vaccines** | 969<br>(27.61) | 2541<br>(72.39) | 337<br>(9.60) | 3173<br>(90.40) | 96<br>(2.74) | 3414<br>(97.26) | 1814<br>(51.68) | 1696<br>(48.32) | 1521<br>(43.33) | 1989<br>(56.67) |
| *P-value* | *0.016* |  | *<0.001* |  | *<0.001* |  | *<0.001* |  | *<0.001* |  |
| **Best COVID vaccine** |  |  |  |  |  |  |  |  |  |  |
| **Moderna** | 10<br>(41.67) | 14<br>(58.33) | 2<br>(8.33) | 22<br>(91.67) | 0 (0) | 24<br>(100) | 10<br>(41.67) | 14<br>(58.33) | 13<br>(54.17) | 11<br>(45.83) |
| **Pfizer- BioNTech** | 69<br>(35.57) | 125<br>(64.43) | 13<br>(6.70) | 181<br>(93.30) | 3<br>(1.55) | 191<br>(98.45) | 117<br>(60.31) | 77<br>(39.69) | 94<br>(48.45) | 100<br>(51.55) |
| **Oxford-AstraZeneca** | 8<br>(15.38) | 44<br>(84.62) | 4<br>(7.69) | 48<br>(92.31) | 1<br>(1.92) | 51<br>(98.08) | 30<br>(57.69) | 22<br>(42.31) | 24<br>(46.15) | 28<br>(53.84) |
| **Sinopharm** | 25<br>(49.02) | 26<br>(50.98) | 7<br>(13.73) | 44<br>(86.27) | 1<br>(1.96) | 50<br>(98.04) | 29<br>(56.86) | 22<br>(43.14) | 24<br>(47.06) | 27<br>(52.94) |
| **Sputnik V** | 0(0) | 9 (100) | 1<br>(11.11) | 8<br>(88.89) | 2<br>(22.22) | 7<br>(77.78) | 4<br>(44.44) | 5<br>(55.56) | 1<br>(11.11) | 8<br>(88.89) |
| *P-value* | *<0.001* |  | *0.596* |  | *0.001* |  | *0.444* |  | *0.256* |  |
| **Getting COVID after vaccination** | 17<br>(35.42) | 31<br>(64.58) | 6<br>(12.50) | 42<br>(87.50) | 1<br>(2.08) | 47<br>(97.92) | 15<br>(31.25) | 33<br>(68.75) | 20<br>(41.67) | 28<br>(58.33) |
| *P-value* | *1* |  | *1* |  | *0.828* |  | *0.008* |  | *0.981* |  |
| **COVID vaccine Instructions** | 64<br>(51.20) | 61<br>(48.80) | 8<br>(6.40) | 117<br>(93.60) | 2<br>(1.60) | 123<br>(98.40) | 61<br>(48.80) | 64<br>(51.20) | 72<br>(57.60) | 53<br>(42.40) |
| *P-value* | *<0.001* |  | *0.003* |  | *0.479* |  | *0.975* |  | *<0.001* |  |
| **Internet search about COVID vaccine** | 60<br>(44.44) | 72<br>(55.65) | 23<br>(17.04) | 112<br>(82.96) | 2<br>(1.48) | 133<br>(98.52) | 71<br>(52.59) | 64<br>(47.41) | 64<br>(47.41) | 71<br>(52.59) |
| *P-value* | *<0.001* |  | *0.02* |  | *0.345* |  | *0.25* |  | *0.119* |  |
| **Getting COVID vaccine if free** | 61<br>(43.57) | 79<br>(56.43) | 18<br>(12.86) | 122<br>(87.14) | 2<br>(1.43) | 138(98.57) | 70(50.00) | 70(50.00) | 71(50.71) | 69(49.29) |
| *P-value* | *<0.001* |  | *1* |  | *0.285* |  | *0.835* |  | *0.003* |  |

#### Confidence

The following variables significantly affected confidence domain: male sex (p<0.001), marital status (p= 0.006), educational level (p= 0.003), previous history of COVID-19 infection (p< 0.001), relatives died due to COVID-19 infection (p= 0.033), taking yearly Flu vaccine (p= 0.007), knowing about different types of vaccine (p= 0.016), following COVID-19 protective measures (p< 0.001), internet search for COVID-19 related information (p< 0.001) and cost of the vaccine (p< 0.001).

#### Complacency

Educational level (p= 0.011), being HCP (p<0.001), previous history of COVID-19 infection (p= 0.034), knowing about different types of vaccine (p< 0.001), following COVID-19 protective measures (p= 0.003), and internet search for COVID-19 related information (p= 0.02) significantly predicted complacency among participants.

#### Constraints

The COVID-19 related constraint was significantly affected by being HCP (p= 0.004), previously infected with COVID-19 (p= 0.001), and knowing about different types of vaccines (p< 0.001).

#### Calculation

The calculation domain was significantly affected by sex (p= 0.007), marital status (p= 0.011), educational level (p< 0.001), being HCP (p< 0.001), having at least one relative died due to COVID-19 (p= 0.002), knowing about the different available types of vaccine (p< 0.001), and believing that there was a risk to get COVID-19 even after vaccination (p= 0.008).

#### Collective responsibility

Educational level (p< 0.001), working as HCP (p< 0.001), previously infected with COVID-19 (p< 0.001), receiving Influenza vaccine yearly (p= 0.049), knowing about the different types of COVID-19 vaccine (p< 0.001), following COVID-19 protective measures (p< 0.001), and availability of vaccine for free (p= 0.003) affected the collective responsibility domain.

### Determinants of phycological antecedents of vaccination

Table 3 shows the regression analysis of the predictors affecting the psychological antecedents. Male sex (OR= 1.428) (95% CI: 1.206 - 1.690), age (OR= 1.029) (95% CI: 1.019 - 1.040), pre-university education (OR= 1.991) (95% CI: 1.249 -3.173), previously infected with COVID-19 (OR= 1.801) (95% CI: 1.458 -2.224), relative infected with COVID-19 (OR= 0.791) (95% CI 0.662 - 0.946) and relative died from COVID-19 (OR= 1.245) (95% CI 1.038 - 1.495) significantly predicted the confidence antecedent.

**Table 3:** Predictors of the 5C Domains

| Independent variables | Odd Ratio | P-value | 95% C.I. for Odd Ratio |
| --- | --- | --- | --- |
| **Confidence** |  |  |  |
| Constant | .065 | <0.001* |  |
| **Sex (Male)** <sup>a</sup> | 1.428 | <0.001* | 1.206 - 1.690 |
| **Age** | 1.029 | <0.001* | 1.019 -1.040 |
| **Education** <sup>b</sup> |  | 0.024* |  |
| Pre-university | 1.991 | 0.004* | 1.249 -3.173 |
| **Previously infected with COVID-19** <sup>c</sup> | 1.801 | <0.001* | 1.458 -2.224 |
| **Relative infected with COVID-19** <sup>d</sup> | .791 | 0.01* | 0.662 - 0.946 |
| **Relatives died from COVID-19** <sup>e</sup> | 1.245 | 0.019* | 1.038 - 1.495 |
| **Complacency** |  |  |  |
| Constant | .144 | <0.001* |  |
| **Education** <sup>b</sup> |  | 0.045* |  |
| University degree | .496 | 0.004* | 0.309 - 0.799 |
| **HCP** <sup>f</sup> | .512 | <0.001* | 0.387 - 0.678 |
| **previously infected with COVID-19** <sup>c</sup> | 1.556 | 0.002* | 1.171 -2.068 |
| **Constraints** |  |  |  |
| Constant | .03 | <0.001* |  |
| **HCP** <sup>f</sup> | .518 | 0.008* | 0.319 -0.841 |
| **Previously infected with COVID-19** <sup>c</sup> | 2.309 | <0.001* | 1.461 -3.649 |
| **Calculation** |  |  |  |
| Constant | .348 | <0.001* |  |
| **Sex (Male)** <sup>a</sup> | 1.362 | <0.001* | 1.169 -1.586 |
| **Age** | 1.012 | 0.014* | 1.002 -1.021 |
| **Education** <sup>b</sup> |  | <0.001* |  |
| University degree | 1.459 | 0.02* | 1.061 -2.007 |
| **HCP** <sup>f</sup> | 1.268 | 0.003* | 1.082 -1.486 |
| **Relatives died from COVID-19** <sup>e</sup> | 1.248 | 0.007* | 1.064 -1.463 |
| **Collective responsibility** |  |  |  |
| Constant | .439 | <0.001* |  |
| **Age** | 1.014 | 0.005* | 1.004 -1.023 |
| **HCP** <sup>f</sup> | 1.594 | <0.001* | 1.358 -1.872 |
| **Previously infected with COVID-19** <sup>c</sup> | .613 | <0.001* | 0.510 -0.736 |
| **Relative infected with COVID-19** <sup>d</sup> | 1.261 | 0.005* | 1.072 -1.482 |

\*, Statistically significant, <sup>a</sup> ref; Female, <sup>b</sup> ref; other education, <sup>c</sup> ref; Not getting COVID, <sup>d</sup> ref: relative not getting COVID, <sup>e</sup> ref; no relative died, <sup>f</sup> ref; No HCP

The significant predictors of complacency are: having a university degree (OR= 0.496) (95% CI 0.309 - 0.799), being a HCP (OR= 0.512) (95% CI: 0.387 - 0.678) and previously infected with COVID-19 (OR= 1.556) (95% CI: 1.171 - 2.068), while the significant predictors of constraints are being a HCP (OR= 0.518) (95% CI 0.319 - 0.841) and infected with COVID-19 before (OR= 2.309) (95% CI 1.461 -3.649).

Male sex (OR= 1.362) (95% CI 1.169 -1.586), age (OR= 1.012) (95% CI 1.002 -1.021), having a university degree (OR= 1.459) (95% CI 1.061 -2.007), being a HCP (OR= 1.268) (95% CI 1.082 -1.486), and having a relative died from COVID-19 (OR= 1.248) (95% CI 1.064 - 1.463) are the significant predictors of calculation among participants, while the significant predictors of collective responsibility are: age (OR= 1.014) (95% CI 1.004 -1.023), being a HCP (OR= 1.594) (95% CI 1.358 -1.872), getting COVID-19 infection before (OR= 0.613) (95% CI 0.510 -0.736) and having a relative infected with COVID-19 (OR= 1.261) (95% CI 1.072 - 1.482).

## Discussion

To understand the reasons behind poor vaccination uptake and low acceptance, tools such as the 5C aim to determine individuals’ psychological antecedents towards vaccination and to design interventions and develop appropriate actions. ^(17)^ This study was able to determine psychological antecedents of vaccine acceptance and determinants of hesitancy within a large number of Arab countries. This large multinational study aimed to assess COVID-19 vaccines psychological antecedent using an online disseminated validated Arabic version of the 5C scale.^(24)^ A total of 4,475 responses from 13 Arab countries were collected. Overall, for every ten participants, about 3 were confident to receive the vaccine, nine showed no complacency toward the COVID-19 vaccine, 5 had calculations, and 4 demonstrated collective responsibility toward the COVID-19 vaccine. Only 3.5% exhibited constraints against COVID-19 vaccination.

The 5C psychological antecedents were previously developed in German and English to measure vaccines acceptance determinants. ^(17)^ Then a protocol on how to culturally adapted and use it with other populations and groups was developed by the same group. ^(27)^ Ghazy et al (2021) were able to demonstrate that it has discriminatory satisfactory power to predict the psychological antecedents of COVID-19 vaccine acceptance and were able to create a cut-off score used in this study. ^(25)^ In this study, it was found that the highest confidence was among the population from UAE, Saudi Arabia, and Kuwait, while the lowest confidence was among the population from Egypt. This finding is supported by the fact that most of the population in UAE and Saudi Arabia had been vaccinated with at least one dose of COVID-19 vaccination, while on the other hand, Egypt’s data regarding vaccination of population is still unclear but in general, less than 4% of the population had been vaccinated. Four vaccines were approved to be used in UAE; Sinopharm, Pfizer-BioNTech, Sputnik V, and Oxford-AstraZeneca, while Saudi Arabia approved three vaccines; Pfizer-BioNTech, Oxford-AstraZeneca, and Moderna vaccines. On the other hand, only two COVID-19 vaccines were used in Egypt; Sinopharm and Oxford-AstraZeneca. The discrepancy in the confidence among the population reflects the population confidence in COVID-19 vaccination itself and its authorities. Cultural differences between Gulf countries and other Arab countries had a role in the confidence of the population. Another effect on confidence is the role of public education and awareness efforts targeting precautions and how to reduce infection and the importance of vaccination. ^(28)^ A recent study by Al-Sanafi & Sallam (2021) showed that Health care workers in Kuwait showed a high intention rate to receive vaccination and that may be again due to the high number of cases in the population, availability of vaccines, and educational efforts, and policies imposed by authorities. ^(29)^ The similarity in Poland and Canada HCPs demonstrated a high acceptance rate. ^(30, 31)^

Complacency was higher among the Moroccan and Jordan populations, while the lowest was among the population from Lebanon. Complacent people see that the vaccination is not important as their immune system can protect them from being infected with COVID-19. Chinese people saw themselves as having good health which in turn affects their intention to be vaccinated. ^(32)^

Similarly for constraints which were higher among Moroccan and Iraqi population. They had their own psychological barriers to avoid taking the vaccine although Morocco had vaccinated nearly 10 million of their population and 12% of them were fully vaccinated, and Morocco ranked the first country in Africa vaccinating its population as it received more vaccination doses than any other African country and designed a large number of vaccination centers and mobile vaccination teams for vaccinating its population. ^(33, 34)^ Since the vaccination against COVID-19 started, Morocco’s government launched communication campaigns to offer information, reassurance and raise the courage of people to be vaccinated. ^(34)^ The main barrier of Moroccan people is the European refusal of Sinopharm and Sputnik V vaccines which restrict their travel to Europe. ^(35)^

The highest calculation score was among Sudanese and Egyptians. Weighing between benefits and risks of being vaccinated before taking the decision. Both Sudan and Egypt did not reach one million people vaccinated up today (140 and 660 thousand in Sudan and Egypt, respectively). ^(33)^ Access to the right information about vaccines through local media and authorities, as well as limited awareness campaigns and lack of defined strategy and transparency of government for vaccine distribution, maybe a possible reason for the low level of calculation among these populations. ^(36)^

The Emiratis had a higher collective responsibility regarding the COVID-19 vaccine. They are willing to protect themselves and others from being infected with the COVID-19 virus. Emirati population as well residents are among the highest populations in the Arab world to encourage social responsibility and collective responsibility. For the last few years, the UAE has been embedding these concepts through educational and social programs. The UAE national charter envisioned the importance of social commitment and responsibility through its mission and vision. ^(37)^ The UAE 2030 agenda for Sustainable Development Goals (SDGs) incorporated its goal to improve solidarity and unity as well as social responsibility. ^(38)^

Predictors affecting the five psychological antecedents vary between these antecedents, however, they are mainly related to being male, of advanced age, education, being a HCP, getting COVID-19, or having a relative infected or died from COVID-19. A similar study conducted among Arab countries found that gender, academic background, attitude towards flu vaccine, infection with COVID-19, and knowledge regarding the type of vaccine had a correlation with vaccine acceptance. ^(39)^ The results from Turkey and United Kingdom (UK) say that male, high education degree, and having children affect vaccine acceptance. ^(40)^ Also, results from low- and middle-income countries showed the willingness of persons to be protected from COVID-19 was the main reason for their acceptance to be vaccinated, while the fear from side effects was the main cause of VH. ^(23)^

Sallam et al (2021) ^(41)^ found an acceptance rate of COVID-19 vaccine of only 29% of their participants which showed a low acceptance rate worldwide compared to acceptance rate ranged from 55% to 90% in other countries. ^(42-44)^ American Study concluded that their high VH contributed to the beliefs that political and social factors and pressures were behind the accelerated approval of COVID-19 vaccines before complete testing of their efficacy and safety. ^(45)^ At the same time, a systematic review found variability of COVID-19 vaccination acceptance between countries with a large number having an acceptance rate of less than 60% which reflects the challenge in controlling the COVID-19 pandemic. The lower rate was reported mainly from Middle East, Russia, and Eastern Europe, while higher rates were reported from East and Southeast Asia. It concluded that COVID-19 VH has a major role in controlling the pandemic which in turn needs a collaborative response from governments, policymakers, and media. ^(46)^ Another meta-analysis found a VH of 17% and a pooled vaccine acceptance of 75%. It reported two reasons determining vaccine acceptance which are case fatality and the number of COVID-19 cases, while the most powerful cause affecting intention to be vaccinated was the people’s trust in the safety of vaccines provided by their countries. ^(47)^

Soares et al (2021) studied the determinants of vaccine hesitancy and found that young age, loss of income during the pandemic, no intention of taking the flu vaccine, low confidence in health care systems during the pandemic, perception of adequacy measures taken by the governments, inadequate information announced by health authorities and low confidence in COVID-19 vaccine safety and efficacy were the factors affecting the refusal or delay of taking COVID-19 vaccine. ^(48)^ Studies around the world have identified reasons and prevalence of COVID-19 VH. In the Arab world, few studies were conducted and pointed out COVID-19 VH. In a recent pre -COVID-19 study conducted in the UAE, 12% of parents were vaccine-hesitant with concerns related to side effects, safety, and multiple injection sites. ^(49)^ In another qualitative study, health care professionals in the UAE showed some hesitancy and training needs on vaccine hesitancy. ^(50)^ Many recent studies have investigated COVID-19 vaccines acceptance around the world. ^(13, 49, 50)^ In Kuwait, a large number of HCPs had a satisfied acceptance of the COVID-19 vaccine with VH concentrated more among female HCPs, nurses, and HCPs working in private facilities, ^(29)^ while in Egypt, the level of VH among medical students was reported at 46% with the main concerns were side effects and ineffectiveness of COVID-19 vaccine. ^(51)^ In Jordan, willingness to take the COVID-19 vaccine (acceptance rates) with a significant portion of the population being indecisive. Participants were reported to be equally distributed between being willing, unwilling, and indecisive to take the COVID-19 vaccine. Participants reported more COVID-19 acceptance for the elderly than themselves. Vaccine development and availability are necessary but not sufficient to achieve immunity against a disease such as COVID-19. Reducing the incidence and prevalence of COVID-19, therefore, necessitates high acceptance and coverage rates to ensure high rates of the population will receive the vaccine. ^(52-55)^

### Strength and Limitations

One of the major strengths of the present study is the large sample size, in addition, the survey population is diverse with representation from different countries, age groups, ethnic and cultural backgrounds. However, the use of convenience sampling, online distribution of the study tool, and sample size determination which did not follow the standard procedure would limit the generalization of the study results to the region of interest. furthermore, there is a risk of selection bias favoring only individuals with access to the internet. A self-reported questionnaire was used to collect the data; hence our findings may be affected by social desirability bias. Despite the stated limitations, the study findings are not inconsistent with previous studies that reported the behavioral factors associated with COVID-19 vaccination hesitancy. More importantly, the study was able to shed light on the overlooked psychological antecedents of COVID-19 vaccination behavior in Arab countries for better policies and actions.

## Conclusion and Recommendations

The study found wide variations in the psychological antecedents of COVID-19 vaccination between the studied countries. Confidence and collective responsibility were higher in countries with high vaccination rates and lower in countries with low vaccination rates. However, other psychological parameters (complacency, constraints, and calculation) differed across countries with varying vaccination rates. Gender, education, getting infected, or having a relative infected or died were the predictors affecting the five psychological antecedents of vaccination. Governments’ decisions and policies, media, and health care authorities must have a role in changing the behavior of the population toward COVID-19 vaccines to insure optimal vaccine acceptance in the region.

## Data Availability

all data are available in the manuscript

## Author contribution

- **Marwa Shawky Abdou:** Conceptualization of research idea, data collection and drafting, and writing and reviewing the manuscript.
- **Ramy Mohamed Ghazy:** Conceptualization of research idea, data collection, and writing the manuscript.
- **Khalid A. Kheriallah:** Assisted in data collection, writing, and revising the manuscript.
- **Maged Ossama Aly:** Assisted in data collection, writing, and revising the manuscript.
- **Ahmed Ramadan:** Performed the statistical analysis, data interpretation, and visualization, and revising the manuscript
- **Yasir Ahmed Mohammed Elhadi:** Assisted in data collection, writing, reviewing, and editing the manuscript.
- **Iffat Elbarazi:** Assisted in data collection, writing, and revising the manuscript.
- **Ehsan Akram Deghidy:** Assisted in data collection and revising the manuscript
- **Haider M. El Saeh:** Assisted in data collection, writing, and revising the manuscript.
- **Karem Mohamed Salem:** Assisted in data collection, writing, and revising the manuscript.

## List of contributors

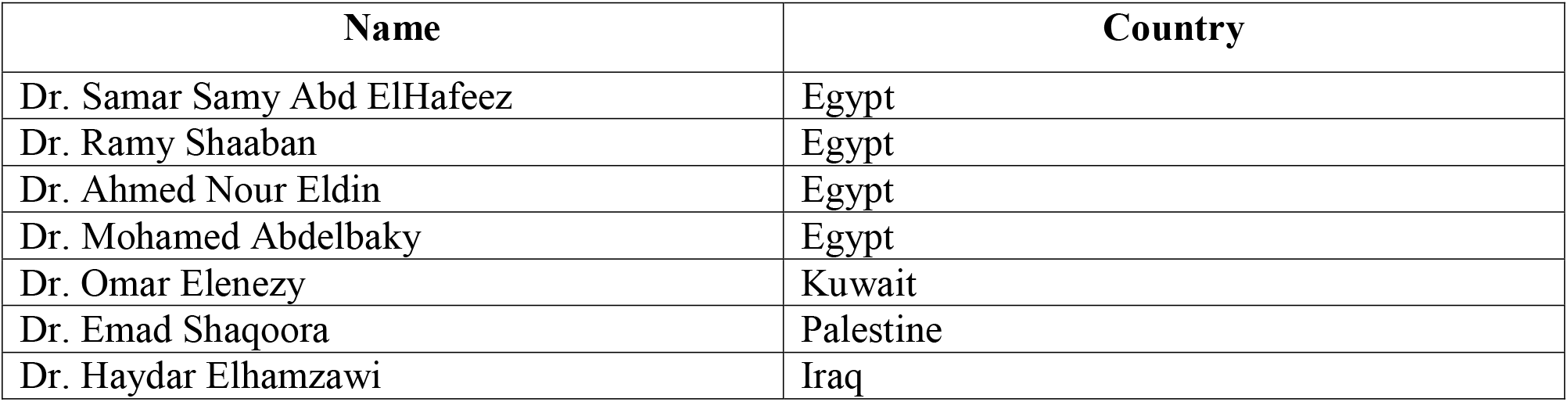
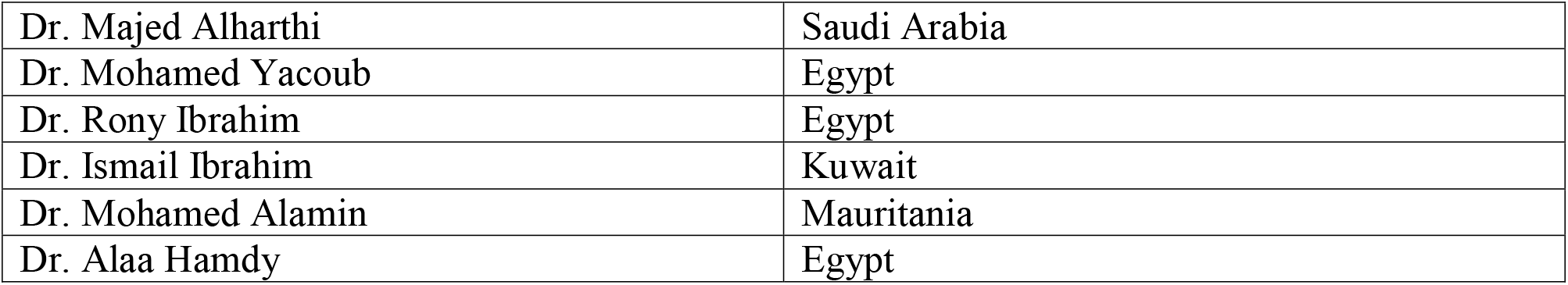

